# CorSeg-CineSAX: An Open-Source Deep Learning Framework for Fully Automatic Segmentation of Short-Axis Cine Cardiac MRI Across Multiple Cardiac Diseases

**DOI:** 10.64898/2026.04.01.26349955

**Authors:** Runhao Xu, Siyuan Jiang, Yujia Zhai, Yuanwei Xu, Yangjie Li, Jie Wang, Lutong Pu, Ke Wan, Shichu Liang, Yucheng Chen

**Affiliations:** Department of Cardiology, West China Hospital, Sichuan University, Chengdu, Sichuan, China; Cardiac Imaging and Target Therapy Lab, West China Hospital, Sichuan University, Chengdu, Sichuan, China

## Abstract

**Background:** Segmentation of the left ventricular (LV) myocardium, LV cavity, and right ventricular (RV) cavity on short-axis cine cardiac magnetic resonance (CMR) images is essential for deriving clinical functional parameters, but existing automated tools rarely combine full public availability (source code, trained weights, and a deployment tool) with validation across diverse centers, vendors, and disease categories, particularly at the technically demanding basal right/left ventricular outflow tract (RVOT/LVOT) level.

**Methods:** We trained a two-dimensional, slice-by-slice MedNeXt-L model, without region-of-interest cropping, on a private cohort of 2,382 subjects (5 disease categories) from 12 domestic centers, and evaluated it on an independent internal test set (n = 650) and three independent external public datasets (ACDC, M&Ms1, M&Ms2; n = 855 combined), spanning 16 disease categories in total (11 never seen during training). Segmentation accuracy (Dice similarity coefficient [DSC], 95th-percentile Hausdorff distance) was assessed overall, by anatomical slice position (including the basal RVOT/LVOT level), and by disease category, center, vendor, and field strength, alongside agreement between automated and manual clinical functional parameters and benchmarking against human inter-observer variability (three independent observers, 50-subject subsample).

**Results:** Mean DSC was 0.908 on the internal test set and 0.883 on the external test set (combined, 0.893; n = 1,505, 31,440 slices). The basal RVOT/LVOT level achieved a mean DSC of 0.904, comparable to the basal (0.910) and mid-ventricular (0.895) levels; the apex was the most challenging position (mean DSC 0.842). Performance on the 11 disease categories absent from training (mean DSC range, 0.874–0.904) was comparable to that on the five categories represented during training (0.881–0.936). Automated LV cavity and right ventricular segmentation matched or exceeded human inter-observer agreement (DSC 0.923 vs. 0.894 and 0.910 vs. 0.906, respectively), while LV myocardium remained somewhat below it (0.847 vs. 0.875). Agreement with manual functional-parameter measurements was strong for LV/RV volumes and LV mass (intraclass correlation coefficient [ICC] ≥ 0.961) and weaker for LV ejection fraction (ICC 0.855) and RV ejection fraction (ICC 0.900), both below the corresponding human inter-observer benchmarks (0.969 and 0.931).

**Conclusions:** CorSeg-CineSAX provides an openly released (code, trained weights, and a standalone GUI application), transparently validated framework for fully automatic CMR short-axis segmentation, with performance at the basal outflow-tract level comparable to other anatomical positions and segmentation accuracy for the LV cavity and right ventricle approaching human inter-observer variability.

## Introduction

Cardiovascular magnetic resonance (CMR) cine imaging is the reference standard for quantifying ventricular volumes and function, from which clinicians derive indices such as ventricular volumes, ejection fraction, and myocardial mass by delineating ventricular structures at end-diastole (ED) and end-systole (ES)^1^. However, manual contouring of a complete short-axis (SAX) stack is time-consuming (typically 15–30 minutes per exam) and subject to substantial inter- and intra-observer variability^2^. Automated, standardized, and reproducible segmentation is therefore essential for translating CMR’s diagnostic potential into routine clinical practice^1^.

Over the past decade, deep learning has driven SAX ventricular segmentation accuracy close to the level of human inter-observer variability^3^, yet this pursuit of accuracy has not translated into clinical readiness, for two reasons. First, a usability gap: most published models release code without trained weights, or require programming expertise and command-line environments inaccessible to clinical practitioners^4^; conversely, fully-packaged commercial platforms such as cvi42 and SuiteHEART^5^ offer complete, user-friendly functionality but release neither code nor weights, precluding reproducibility and further development. Second, a validation gap: even open-source models are rarely subjected to the cross-center, cross-vendor, and cross-disease testing needed to establish real-world reliability^4^, and are seldom evaluated at the basal right/left ventricular outflow tract (RVOT/LVOT) level—an anatomically demanding region, owing to valve-plane motion through the imaging plane, that current models still do not handle adequately^6^. To our knowledge, no existing open-source model closes both gaps at once.

In this work, we present CorSeg-CineSAX, a fully automated deep learning framework designed to close both gaps at once. Architecturally, CorSeg-CineSAX employs a purely 2D, per-slice MedNeXt-L^7^ backbone trained in a whole-image-in, whole-image-out mode without region-of-interest(ROI) cropping, and we publicly release the source code, trained weights, and a standalone GUI application requiring no programming expertise. We train and validate the model on 3,237 subjects from 18 centers, four vendor platforms, and 14 disease categories in addition to healthy controls, reporting performance transparently by anatomical slice position, including the basal RVOT/LVOT level, and benchmarked directly against human inter-observer variability.

## Methods

### Study Overview

This study aimed to develop and validate an open-source, ready-to-use deep learning tool for fully automatic segmentation of short-axis cine CMR images. The workflow comprised three stages (Figure 1): dataset construction restricted to the end-diastolic (ED) and end-systolic (ES) frames; training of a MedNeXt-L segmentation model in a two-dimensional, slice-by-slice strategy; and comprehensive validation on an internal test set and three external public datasets, including accuracy by anatomical position and disease subgroup, functional-parameter agreement, and benchmarking against human inter-observer variability. Code, weights, and a standalone GUI application are publicly released at https://github.com/RunhaoXu2003/CorSeg.

**Figure 1.**
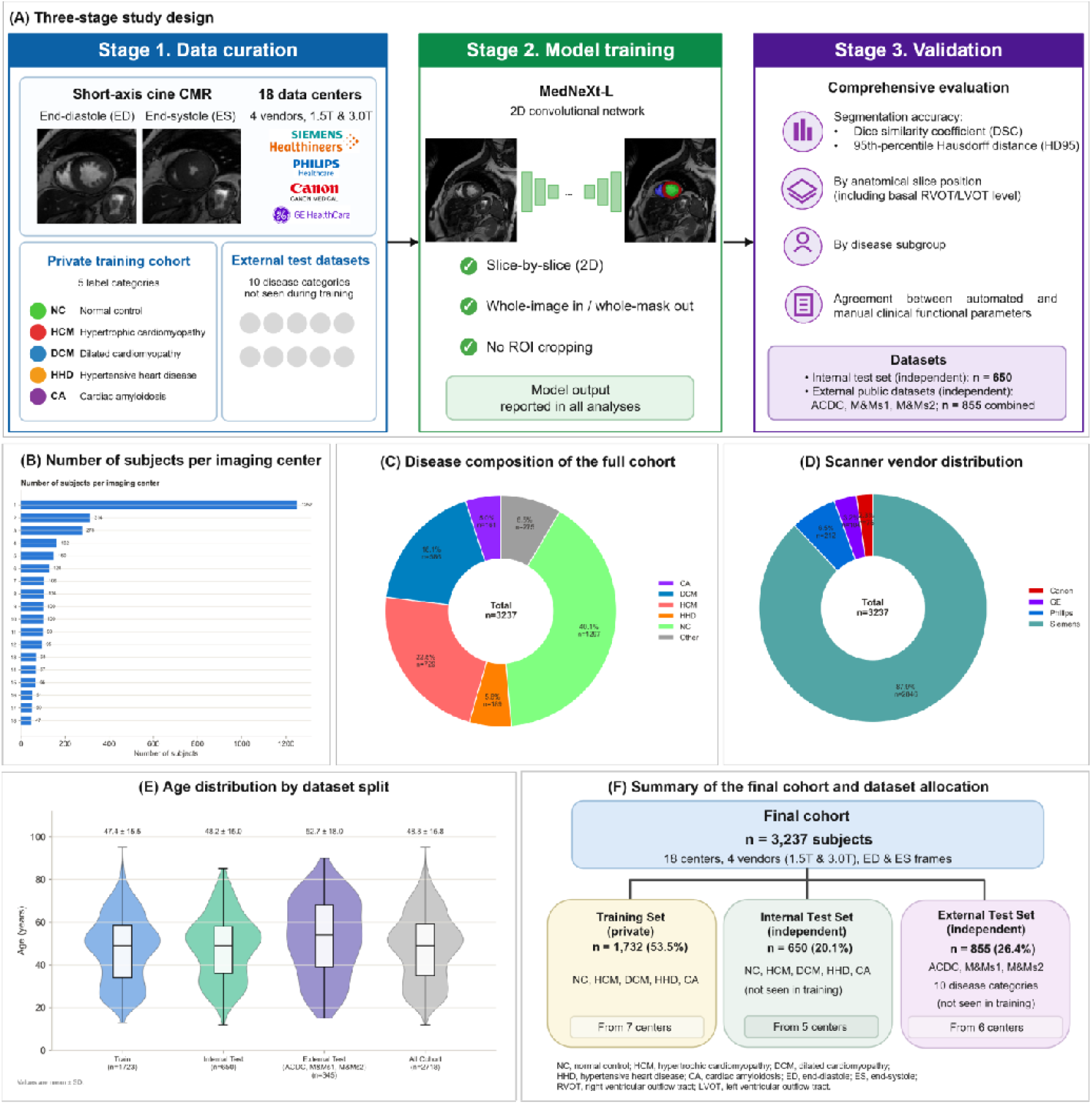
Overview of the CorSeg-CineSAX study workflow and study cohort. (A) Three-stage workflow. **Stage 1 (Data curation):** short-axis cine CMR (ED/ES) from 18 centers, 4 vendors, 1.5/3.0 T; the training cohort covered five disease categories (NC, HCM, DCM, HHD, CA), and the external test sets added ten further categories unseen in training. **Stage 2 (Model training):** MedNeXt-L, 2D, slice-by-slice, whole-image input/output, no ROI cropping. **Stage 3 (Validation):** DSC/HD95 evaluated by slice position (including the basal RVOT/LVOT level) and disease subgroup, plus functional-parameter agreement, on an internal test set (n = 650) and three external public datasets (ACDC, M&Ms1, M&Ms2; n = 855). (B) Subjects per center (18 centers; range, 47–1252). (C) Disease composition (n = 3237): NC 1297 (40.1%), HCM 729 (22.5%), DCM 586 (18.1%), HHD 189 (5.8%), CA 161 (5.0%), Other 275 (8.5%; categories absent from training). (D) Vendor distribution: Siemens 87.9%, Philips 6.5%, GE 3.2%, Canon 2.3%. (E) Age distribution by split (violin/box plots; subjects with recorded age only, hence a smaller n for the external test set than in panel F). (F) Final allocation: training/validation n = 1732 (53.5%), internal test n = 650 (20.1%), external test n = 855 (26.4%; ACDC, M&Ms1, M&Ms2).

### Dataset Collection and Cohort Composition

A total of 2,382 subjects were enrolled from 12 domestic centers, contributing 1,732 subjects to training/validation and 650 to the internal test set (Table 1), across five conditions: normal control (NC), hypertrophic cardiomyopathy (HCM), dilated cardiomyopathy (DCM), hypertensive heart disease (HHD), and cardiac amyloidosis (CA).

**Table 1.** Baseline characteristics of the study cohort, stratified by data split.

| Characteristic | Train | InternalTest | ExternalTest |
| --- | --- | --- | --- |
| N | 1732 | 650 | 855 |
| Age (y), mean±SD | 47.4±15.5 | 48.2±15.0 | 52.7±18.0 |
| Male, n (%) | 1004 (58%) | 380 (58%) | 211 (25%) |
| Female, n (%) | 728 (42%) | 270 (42%) | 134 (16%) |
| Disease CA | 89 | 72 | 0 |
| Disease DCM | 399 | 0 | 187 |
| Disease HCM | 440 | 114 | 175 |
| Disease HHD | 73 | 92 | 24 |
| Disease NC | 731 | 372 | 194 |
| Disease Other | 0 | 0 | 275 |
| Vendor Siemens | 1732 | 650 | 464 |
| Vendor Philips | 0 | 0 | 212 |
| Vendor GE | 0 | 0 | 104 |
| Vendor Canon | 0 | 0 | 75 |
| Field 3.0T | 1732 | 650 | 112 |
| Field 1.5T | 0 | 0 | 593 |
| Field 1.5/3.0T | 0 | 0 | 150 |
| Protocol bSSFP | 1732 | 650 | 0 |
| Protocol SSFP | 0 | 0 | 150 |
| Protocol NR | 0 | 0 | 705 |
| Scanner MAGNETOM Prisma/Skyra | 1732 | 650 | 0 |
| Scanner Achieva | 0 | 0 | 212 |
| Scanner SymphonyTim | 0 | 0 | 151 |
| Scanner MAGNETOM Area / Trio Tim | 0 | 0 | 150 |
| Scanner MAGNETOM Skyra | 0 | 0 | 95 |
| Scanner Vantage Orian | 0 | 0 | 75 |
| Scanner Other | 0 | 0 | 172 |

Values are n, mean ± SD, or n (%). Train = private, single-vendor 3.0 T cohort (West China Hospital); Internal Test = independent private-cohort subjects; External Test = ACDC, M&Ms1, M&Ms2 (none seen in training). “Disease Other” (n = 275) aggregates ten additional disease categories plus a residual group unseen in training (full breakdown, Table S1). ACDC field strength is reported at the dataset level (per-subject values unavailable). NR, protocol not reported.

The NC sub-cohort (n = 1,103) was recruited across 11 domestic centers between September 2019 and May 2022 as part of a previously published reference-value study^8^ (ChiCTR1900025518). The HCM, DCM, HHD, and CA sub-cohorts were contributed by West China Hospital (training/validation) and The First Affiliated Hospital of Anhui Medical University (internal test), following diagnostic criteria from related institutional cohort studies^9^: HCM, wall thickness ≥15 mm (≥13 mm for a first-degree relative) without another cause of hypertrophy; DCM, LV end-diastolic dimension >55/>50 mm (men/women) with LVEF <50%, excluding ischemic, valvular, hypertensive, and hypertrophic causes; HHD, wall thickness ≥12 mm with hypertension and no other cause of hypertrophy; CA (AL subtype), Congo-red–positive biopsy with light-chain confirmation. Recruitment extended from each sub-cohort’s start date (HCM, 2013; DCM, 2012; HHD, 2014; CA, 2011) through 2025.

Images were acquired on 3.0-T Siemens scanners (MAGNETOM Prisma/Skyra) using balanced steady-state free precession sequences, contiguous short-axis slices base-to-apex (slice thickness, 8 mm); only ED and ES frames were annotated and used.

Three public datasets (855 subjects) were used exclusively for external validation. ACDC^10^ (150 subjects, single center) spans five groups (normal, myocardial infarction, dilated/hypertrophic cardiomyopathy, abnormal right ventricle). M&Ms1^11^ (345 subjects, five centers, four vendors) additionally includes hypertensive heart disease, athlete heart syndrome, ischemic heart disease, and left ventricular non-compaction. M&Ms2^6^ (360 subjects, three centers/vendors) adds right-ventricular pathologies (arrhythmogenic cardiomyopathy, Tetralogy of Fallot, tricuspid regurgitation, inter-atrial communication, dilated right ventricle). Label encodings were mapped to a common convention (0 = background, 1 = LV myocardium, 2 = LV cavity, 3 = RV).

Subjects were allocated to training/validation, internal test, or external test at the subject level, with all slices/phases of a subject in one partition. The public datasets served entirely as external test. The Anhui HCM/HHD/CA sub-cohort was assigned entirely to internal test, and the West China CA/DCM/HCM/HHD sub-cohorts entirely to training/validation. The 11-center NC cohort was split by contributing center (7 centers to training/validation, n = 731; 4 centers to internal test, n = 372), together yielding seven training-only and five internal-test-only domestic centers (Figure 1; full allocation in Table 1).

### Manual Reference Standard and Annotation Workflow

Ground truth for the private cohort was established using 3D Slicer (v5.10.0) by three cardiologists (Observer 1–3), who divided the cohort and each annotated a distinct portion, delineating LV myocardium, LV cavity, and RV cavity. Papillary muscles were included within the cavity blood pool and the myocardial boundary was not interpolated at the basal slice, consistent with ACDC/M&Ms1/M&Ms2 conventions^6,10,11^. Annotators referenced long-axis (2-/4-chamber) views to define the LV base/LVOT^10^ and, for the RV, followed the M&Ms2 protocol^6^: RV volume larger at ED than ES, and the basal (RVOT-encompassing) and apical extent defined via the tricuspid-annulus position and the most apical slice with a detectable cavity.

To assess reference-standard reliability, all three observers independently and blindly re-segmented the same 50-subject subsample (10 per disease category × ED/ES).

### Data Preprocessing

Images were preprocessed identically for training and test data (NIfTI loading, label remapping, resampling to 1.25 × 1.25 mm in-plane resolution, resizing to 224 × 224 pixels, per-slice z-score normalization, and training-only data augmentation), using the full field-of-view without region-of-interest cropping. Full preprocessing and augmentation parameters are provided in Supplementary Methods.

### Model Architecture

A MedNeXt-L^7^ convolutional network was trained in MONAI^12^ to process each short-axis slice independently (two-dimensional, whole-image-in/whole-image-out, four output channels), without 3D volumetric context, temporal sequences, or prior ROI localization. Architectural and configuration details are provided in Supplementary Methods.

### Training Protocol

The model was trained end-to-end using a composite Dice/Focal/cross-entropy loss with the AdamW^13^ optimizer and a warmup-cosine-annealing learning-rate schedule. Full training configuration (loss weighting, epochs, batch size, hardware, and reproducibility settings) is provided in Supplementary Methods.

### Evaluation Metrics

Segmentation accuracy was evaluated using the Dice similarity coefficient (DSC) and 95th-percentile Hausdorff distance (HD95) for each foreground structure on every evaluable slice, summarized as mean ± SD with observed range, for the internal, external, and combined test sets, and stratified by anatomical position and disease category.

Slice position was determined per subject from the end-diastolic LV cavity area regressed against slice index; the slope sign identified the base-to-apex direction. The two most basal slices were designated the basal outflow-tract level (RVOT/LVOT); remaining slices were divided into equal thirds (basal, mid-ventricular, apical).

LV/RV volumes (LVEDV/LVESV/RVEDV/RVESV), ejection fractions (LVEF/RVEF), and LV mass were derived from automated and manual segmentations independently, with ED/ES identified as the frames of maximum/minimum LV cavity volume. Volumes were computed as segmented area × pixel spacing × slice thickness (8 mm private cohort; NIfTI header or nominal 10 mm for external data); EF = (EDV − ESV)/EDV × 100%; LV mass = ED myocardial volume × 1.05 g/mL.

Agreement between automated and manual functional parameters was assessed using Bland-Altman bias/limits of agreement, Pearson’s r, the intraclass correlation coefficient (ICC[A,1])^14^, Lin’s concordance correlation coefficient (CCC)^15^, mean absolute error, median error, interquartile range, and mean percentage error.

Pairwise DSC among the three observers was computed on the 50-subject subsample using the same definition as above; a three-rater ICC(A,1) was used for functional parameters.

Eighteen training/validation subjects underwent repeat CMR as routine follow-up (47 scans; median interval, 376 days; range, 28–1,190 days). Pairwise ICC(A,1) and within-subject coefficient of variation were computed from automated measurements alone, reflecting long-term physiological change rather than short-term reproducibility.

### Statistical Analysis

Metrics are summarized descriptively (mean ± SD, range). Between-subject factors (disease category, center, vendor, field strength, source dataset) were compared using the Kruskal-Wallis test on subject-level mean DSC (averaged across evaluable slices within each subject to avoid pseudo-replication), with Holm-Bonferroni-adjusted p-values and epsilon-squared (ε^2^) effect sizes; internal-versus-external performance by the Mann-Whitney U test; cardiac phase (ED vs. ES) by the Wilcoxon signed-rank test; and anatomical position (four related samples) by the Friedman test. A two-sided p < 0.05 was considered statistically significant; because sample sizes are large, effect sizes or median differences are reported alongside p-values throughout. Analyses were performed in Python (SciPy).

## Results

### Cohort Description

The final analysis cohort comprised 3,237 subjects from 18 imaging centers (12 domestic, 6 international), four scanner vendors, and two field strengths (Table 1, Figure 1): training/validation (n = 1,732), internal test (n = 650), and external test (n = 855; ACDC, M&Ms1, M&Ms2). Sixteen disease categories were represented: five in training (NC, n = 1,297; HCM, n = 729; DCM, n = 586; HHD, n = 189; CA, n = 161) and 11 present only in external testing (n = 275 combined). Mean age was similar in the training/validation and internal test sets (47.4±15.5 and 48.2±15.0 years) and higher in the external test set (52.7±18.0 years). Vendor was heavily skewed toward Siemens (2,846/3,237, 87.9%), entirely within the training/validation and internal test sets.

### Overall Segmentation Performance

Table 2 summarizes segmentation performance. Mean DSC was 0.908 on the internal test set (650 subjects, 12,426 slices; LVM 0.872±0.088, LVC 0.925±0.069, RV 0.927±0.076) and 0.883 on the external test set (855 subjects, 19,014 slices; LVM 0.831±0.074, LVC 0.921±0.077, RV 0.898±0.100); combined, mean DSC was 0.893 (1,505 subjects, 31,440 slices). Internal-test performance was modestly higher than external (median subject-level mean DSC, 0.903 vs. 0.882; Mann-Whitney U test, p = 9.6×10□□□), consistent with greater vendor, field-strength, and disease heterogeneity externally, though the absolute difference was small.

**Table 2.** Segmentation performance (DSC and HD95), overall and versus human inter-observer variability.

| Set | Subjects | Slices | LVM DSC | LVC DSC | RV DSC | mean DSC | HD95 LVM | HD95 LVC | HD95 RV |
| --- | --- | --- | --- | --- | --- | --- | --- | --- | --- |
| InternalTest | 650 | 12426 | 0.872±0.088<br>(0.00–0.99) | 0.925±0.069<br>(0.00–0.99) | 0.927±0.076<br>(0.00–1.00) | 0.908 | 2.64±4.33 | 2.72±4.58 | 2.54±2.18 |
| ExternalTest | 855 | 19014 | 0.831±0.074<br>(0.00–0.96) | 0.921±0.077<br>(0.00–0.98) | 0.898±0.100<br>(0.00–0.98) | 0.883 | 2.83±1.57 | 2.59±1.12 | 3.19±2.98 |
| Overall<br>(internal+external) | 1505 | 31440 | 0.847±0.083<br>(0.00–0.99) | 0.923±0.073<br>(0.00–0.99) | 0.910±0.092<br>(0.00–1.00) | 0.893 | 2.75±3.02 | 2.64±3.06 | 2.92±2.69 |
| Inter-observer<br>(human, n=50, 3<br>raters pairwise) | 50 | 2727 | 0.875±0.084<br>(0.27–0.99) | 0.894±0.049<br>(0.76–1.00) | 0.906±0.048<br>(0.15–0.99) | 0.892 | — | — | — |

P value (not shown in table above): Internal vs. external test set (subject-level mean DSC): Mann-Whitney U test, $U = 397781$ , $P = 9.6 \times 10^{-16}$ (medians 0.903 vs. 0.882).

### Performance by Anatomical Position and Other Sources of Heterogeneity

Segmentation accuracy varied significantly by anatomical slice position (Table 3; Friedman test, χ^2^ = 1696.9, p < 0.001). The basal outflow-tract level (RVOT/LVOT) achieved mean DSC 0.904, comparable to the basal (0.910) and mid-ventricular (0.895) levels; the apex was most challenging (mean DSC 0.842), with accuracy declining progressively from base to apex rather than being lowest at the outflow tract.

**Table 3.** Segmentation performance stratified by anatomical short-axis slice position, including the basal RVOT/LVOT level.

| Position | Subjects | Slices | LVM DSC | LVC DSC | RV DSC | mean DSC | HD95 LVM | HD95 LVC | HD95 RV |
| --- | --- | --- | --- | --- | --- | --- | --- | --- | --- |
| basal_outflow | 1505 | 6108 | 0.852±0.096<br>(0.00–0.98) | 0.930±0.067<br>(0.00–0.99) | 0.929±0.095<br>(0.00–0.99) | 0.904 | 2.76±3.05 | 2.69±3.24 | 3.22±4.72 |
| basal | 1505 | 9504 | 0.859±0.063<br>(0.11–0.99) | 0.939±0.040<br>(0.00–0.99) | 0.931±0.053<br>(0.00–0.99) | 0.910 | 2.75±3.33 | 2.64±2.95 | 2.91±2.45 |
| mid | 1505 | 8484 | 0.851±0.066<br>(0.00–0.99) | 0.928±0.048<br>(0.00–0.99) | 0.906±0.072<br>(0.00–1.00) | 0.895 | 2.72±3.13 | 2.68±3.67 | 2.84±2.13 |
| apical | 1505 | 7344 | 0.814±0.119<br>(0.00–0.98) | 0.867±0.131<br>(0.00–0.98) | 0.843±0.171<br>(0.00–0.99) | 0.842 | 2.80±1.86 | 2.52±1.20 | 2.87±2.34 |

P value (not shown in table above): Comparison across the four anatomical positions (subject-level mean DSC, four related samples): Friedman test, $\chi^2 = 1696.9$ , $P < 0.001$ (n = 1048 subjects with all four positions present).
DSC and HD95 (mm), mean ± SD (min–max) by slice position — basal outflow tract (RVOT/LVOT level), basal, mid-ventricular, apical — combined test set (n = 1505). Position was assigned by linear regression of LV cavity area vs. slice index at end-diastole, which reassigned the basal/apical direction for 24.1% of subjects (0–66% across sub-cohorts) relative to an earlier method.

Performance also varied significantly by disease category, center, vendor, and field strength (Kruskal-Wallis tests, Holm-adjusted p < 10□^2^□; Supplementary Tables S1–S3b), with center showing the largest effect (ε^2^ = 0.53) and vendor the smallest (ε^2^ = 0.09). By disease (Table S1), mean DSC ranged from 0.874 (myocardial infarction with reduced ejection fraction) to 0.936 (cardiac amyloidosis); the 11 categories unseen during training spanned a comparable range (0.874–0.904) to the five trained categories (0.881–0.936). Ischemic heart disease (n = 5) reflects its sole source dataset’s (ACDC) case distribution. By center (Table S2), mean DSC ranged from 0.859 to 0.937; by vendor (Table S3a), 0.874–0.899; by field strength (Table S3b), 0.880–0.905. Cardiac phase also differed significantly (Wilcoxon signed-rank test, p = 1.0×10□^1^□; Table S4), with a small absolute difference (median 0.891 vs. 0.888).

### Qualitative Results

Figure 2 shows qualitative segmentation at the basal RVOT/LVOT and RV levels in disease categories absent from training and across the base-to-apex extent in the private cohort, with close visual agreement between predictions and ground truth. Supplementary Figure S3 additionally presents a normal control alongside two pathological examples at each of the LVOT, RVOT, and RV levels.

**Figure 2.**
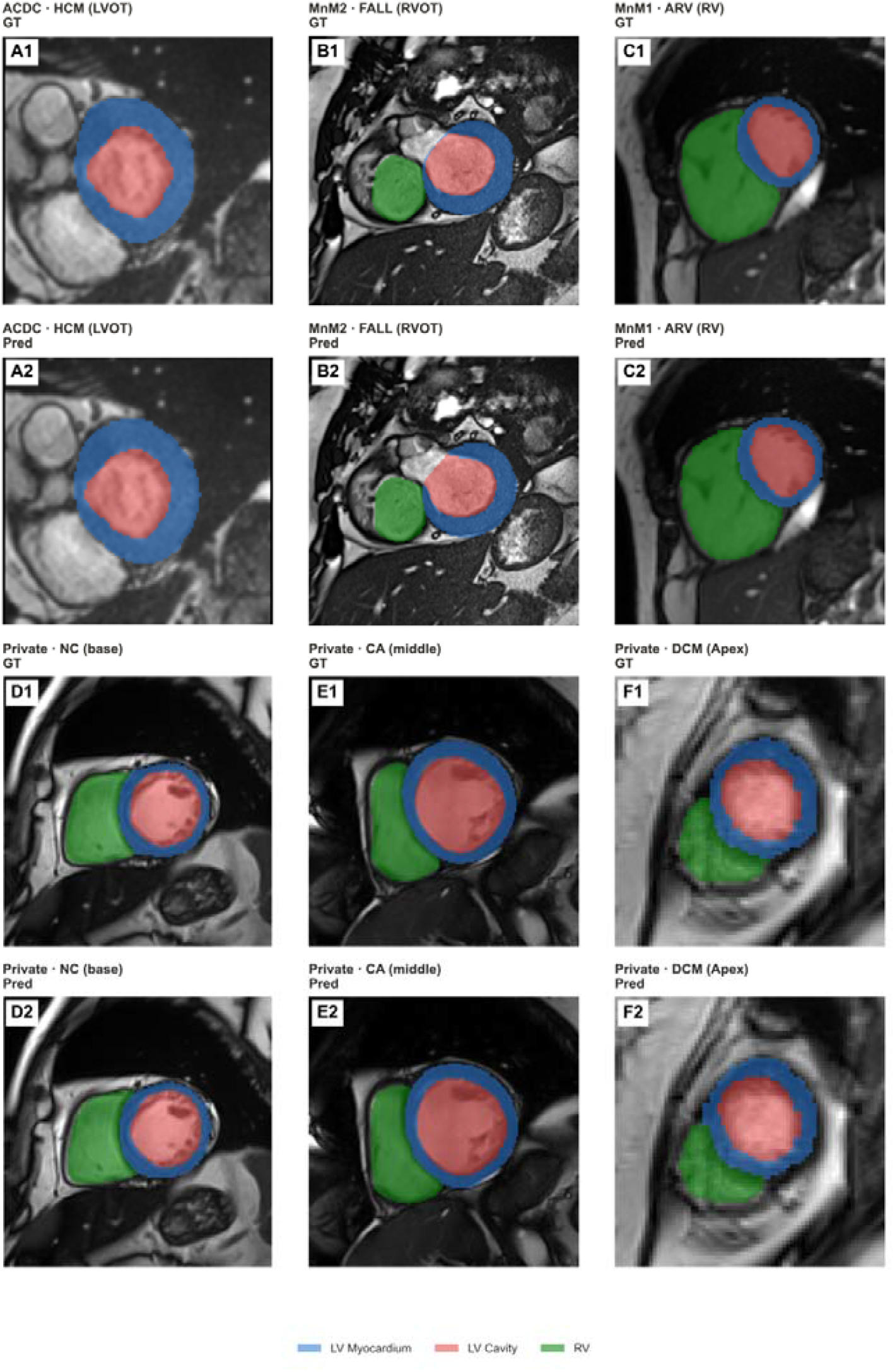
Qualitative segmentation at the basal RVOT/LVOT and right-ventricular levels, and across the base-to-apex extent, in normal and pathological cases. GT (top) and Pred (bottom) short-axis masks (blue, LV myocardium; red, LV cavity; green, RV; solid fill). **Top row:** basal outflow-tract/RV-level cases from external datasets, disease categories unseen in training — **(A)** ACDC, HCM, LVOT level; **(B)** M&Ms2, Tetralogy of Fallot (FALL), RVOT level; **(C)** M&Ms1, abnormal right ventricle (ARV), RV-dominant level. **Bottom row:** base-to-apex examples from the private cohort — **(D)** NC, basal; **(E)** CA, mid-ventricular; **(F)** DCM, apical. Panels labelled “1” are GT; “2” are the corresponding Pred for the same slice.

Figure 3 shows four representative failure cases: non-detection of the LV cavity with a fragmented myocardial prediction in severe hypertrophic cardiomyopathy; non-detection of both the LV cavity and RV at a technically demanding slice; isolated non-detection of the RV in an abnormal-right-ventricle subject; and a false-positive segmentation at an anatomically implausible location.

**Figure 3.**
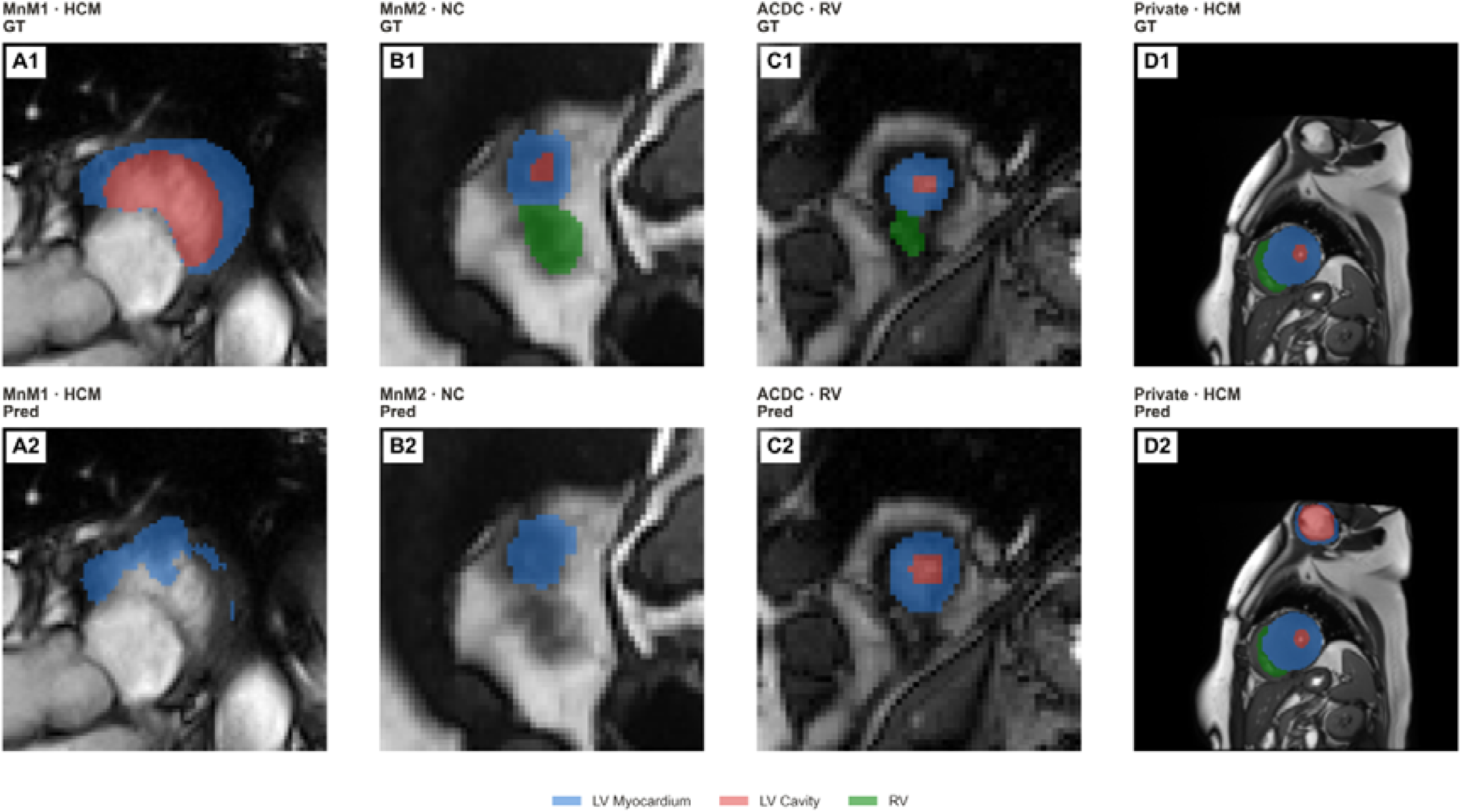
Representative segmentation failure cases. GT (top) and Pred (bottom) masks (blue, LV myocardium; red, LV cavity; green, RV) for four failure modes. **(A)** M&Ms1, HCM: fragmented myocardial contour, LV cavity undetected. **(B)** M&Ms2, NC: LV cavity and RV both missed at a small-structure slice. **(C)** ACDC, abnormal RV: LV segmented correctly, RV missed entirely. **(D)** private cohort, HCM: segmentation was accurate at the true cardiac region, but a false-positive full segmentation appeared at another region with no cardiac substrate in GT — illustrating a risk of whole-image, non-cropped inference.

### Agreement Between Automated and Manual Clinical Functional Parameters

Table 4 and Figure 4 summarize functional-parameter agreement (n = 1,505). LV parameters showed a consistent, modest overestimation bias: LVEDV (+9.35 mL, ICC = 0.974), LVESV (+10.88 mL, ICC = 0.961), LV mass (+6.99 g, ICC = 0.968). LVEF showed weaker agreement (bias −5.40%, r = 0.909, ICC = 0.855; mean absolute error [MAE] 6.20%), with systematic underestimation. RV parameters were comparable to LV for RVEDV (ICC = 0.973) and RVESV (ICC = 0.967); RVEF had the smallest bias of any parameter (−1.40%) with similarly moderate correlation (r = 0.906, ICC = 0.900; MAE 3.97%). Mean percentage error ranged from −8.4% (LVEF) to +28.7% (LVESV).

**Table 4.** Agreement between automated and manual measurements of clinical functional parameters (n = 1505).

| Param | n | GT<br>(mean±SD) | Auto<br>(mean±SD) | Bias | 95%LoA | r | ICC | CCC | MAE | MedErr | IQR | %Err | Inter-observer<br>ICC<br>(human) |
| --- | --- | --- | --- | --- | --- | --- | --- | --- | --- | --- | --- | --- | --- |
| LVEDV | 1505 | 136.1±66.0 | 145.4±67.9 | +9.35 | [-14.63,<br>+33.33] | 0.984 | 0.974 | 0.974 | 10.14 | +5.44 | 13.81 | +8.1 | 0.985 |
| LVESV | 1505 | 62.6±55.4 | 73.5±57.0 | +10.88 | [-11.68,<br>+33.44] | 0.979 | 0.961 | 0.961 | 11.26 | +9.14 | 13.95 | +28.7 | 0.987 |
| LVEF | 1505 | 58.1±16.4 | 52.7±15.0 | -5.40 | [-18.83,<br>+8.03] | 0.909 | 0.855 | 0.855 | 6.20 | -3.71 | 8.88 | -8.4 | 0.969 |
| LVMass | 1505 | 103.8±53.2 | 110.8±53.0 | +6.99 | [-15.78,<br>+29.75] | 0.976 | 0.968 | 0.968 | 10.15 | +6.05 | 13.09 | +8.3 | 0.983 |
| RVEDV | 1505 | 130.2±56.5 | 137.5±59.6 | +7.31 | [-15.20,<br>+29.82] | 0.982 | 0.973 | 0.973 | 8.21 | +3.94 | 8.62 | +6.0 | 0.994 |
| RVESV | 1505 | 66.2±38.5 | 71.5±40.4 | +5.29 | [-11.89,<br>+22.46] | 0.976 | 0.967 | 0.967 | 6.33 | +2.88 | 7.34 | +10.4 | 0.989 |
| RVEF | 1505 | 49.9±14.4 | 48.5±13.7 | -1.40 | [-13.38,<br>+10.58] | 0.906 | 0.900 | 0.900 | 3.97 | -0.35 | 4.48 | -2.0 | 0.931 |

GT, manual reference; Auto, model measurement; Bias, mean auto–manual difference; 95% LoA, bias ± 1.96 SD; r, Pearson correlation; ICC, intraclass correlation (two-way, absolute agreement); CCC, Lin’s concordance correlation; MAE, mean absolute error; MedErr, median signed error; IQR, interquartile range of signed error; %Err, mean percentage error. Rightmost column: three-rater inter-observer ICC (n = 49, Table S7c), as a human-performance benchmark.

**Figure 4.**
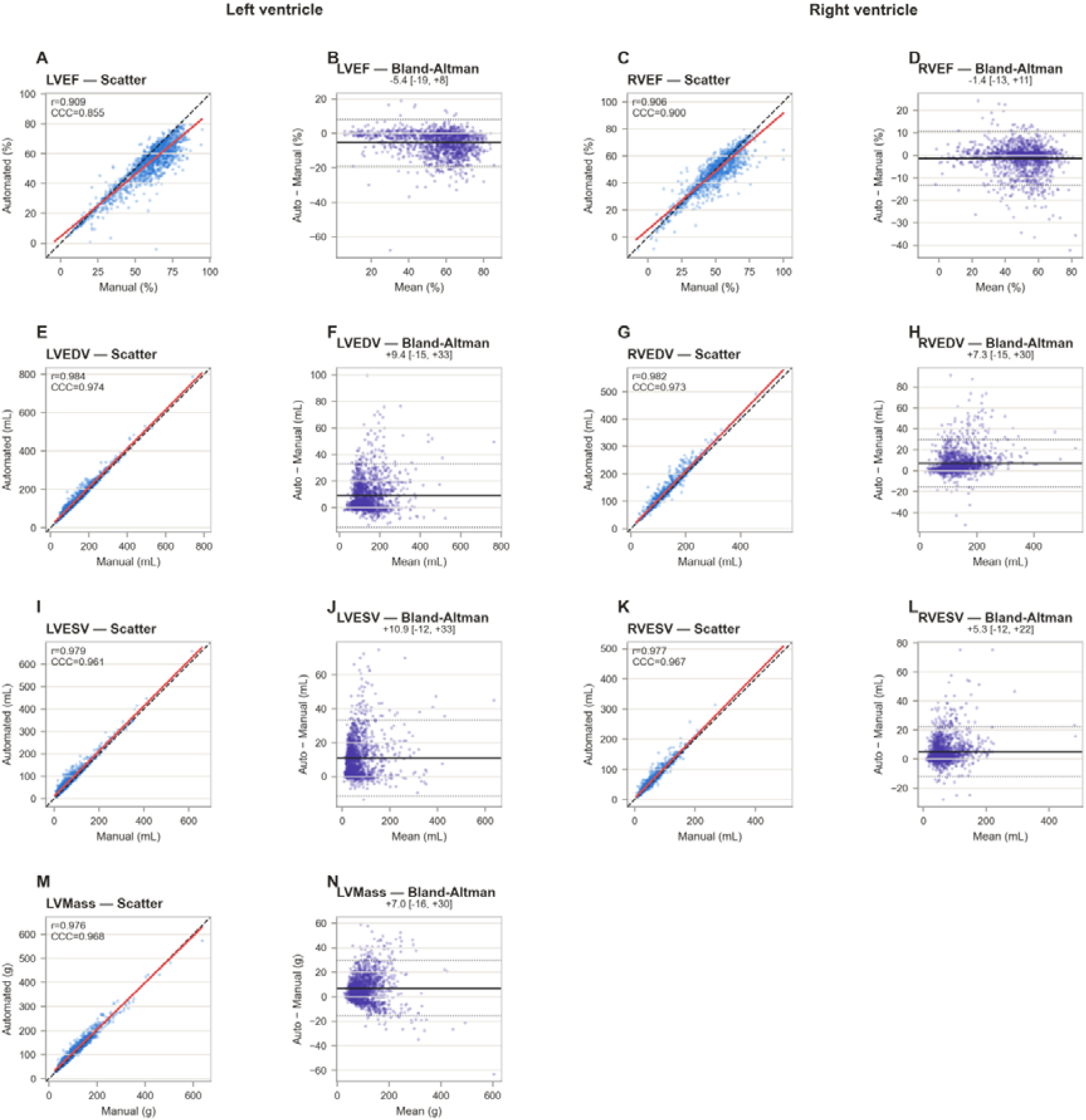
Agreement between automated and manual clinical functional parameters. Scatterplots (identity line, dashed; regression line, red) and Bland-Altman plots for seven functional parameters, automated vs. manual (n = 1505); left columns, LV; right columns, RV. **(A, B) LVEF:** r = 0.909, CCC = 0.855, bias = −5.4% [−18.8, +8.0]. **(C, D) RVEF:** r = 0.906, CCC = 0.900, bias = −1.4% [−13.4, +10.6]. **(E, F) LVEDV:** r = 0.984, CCC = 0.974, bias = +9.4 mL [−14.6, +33.3]. **(G, H) RVEDV:** r = 0.982, CCC = 0.973, bias = +7.3 mL [−15.2, +29.8]. **(I, J) LVESV:** r = 0.979, CCC = 0.961, bias = +10.9 mL [−11.7, +33.4]. **(K, L) RVESV:** r = 0.977, CCC = 0.967, bias = +5.3 mL [−11.9, +22.5]. **(M, N) LVMass:** r = 0.976, CCC = 0.968, bias = +7.0 g [−15.8, +29.8]. Bias, mean auto−manual difference; brackets, 95% limits of agreement.

### Benchmarking Against Human Inter-observer Variability

Pairwise inter-observer DSC (50-subject subsample; Table 2, Table S7) was 0.875 (LVM), 0.894 (LVC), 0.906 (RV; mean 0.892). Automated performance matched or exceeded this for LVC (0.923) and RV (0.910), remaining below it for LVM (0.847). Inter-observer agreement by position (Table S7b) varied little (0.889–0.897), unlike the marked apical decline of the automated model (§3.3), suggesting a genuine model limitation rather than reduced ground-truth reliability there. Three-rater inter-observer ICC for functional parameters (Table S7c) was 0.985– 0.994 (volumes/mass), 0.969 (LVEF), and 0.931 (RVEF)—all higher than the corresponding automated ICC (§3.5).

### Long-Term Repeat-Scan Reproducibility

Among 18 subjects with repeat scans obtained as part of routine clinical follow-up (47 scans; median interval, 376 days; range, 28–1,190 days), automated functional-parameter reproducibility was low to moderate (ICC[A,1], 0.338–0.456; within-subject coefficient of variation, 14.5%–34.7%; Supplementary Table S6), consistent with genuine longitudinal physiological change rather than short-term reproducibility.

## Discussion

### Principal Findings

We developed and validated CorSeg-CineSAX on 3,237 subjects across 18 centers, 4 vendors, and 16 disease categories. The model achieved a mean DSC of 0.908 on the internal test set and 0.883 on the external test set; accuracy at the basal RVOT/LVOT level (0.904) was comparable to other basal and mid-ventricular levels, with the apex proving the most challenging position. Automated LV cavity and right ventricular segmentation matched or exceeded human inter-observer agreement, while LV myocardium and ejection-fraction estimation remained modestly below this benchmark.

### Comparison with Existing Tools and Positioning of This Study’s Contribution

Existing automated CMR segmentation tools rarely combine full public availability with broad, diverse validation: many release code without trained weights^16^ or require programming expertise^4^ or 3D/4D input with manual localization^17^ that real-world acquisitions cannot always guarantee, while commercial platforms such as cvi42 and SuiteHEART^5^ release neither code nor weights. CorSeg-CineSAX processes a single 2D slice with no cropping and releases source code, trained weights, and a standalone application together.

A comparable large-scale study trained on 1,923 scans from 13 institutions and 10 scanner models across 9 conditions^18^; the present cohort is larger in subject and disease-category count, though narrower in vendor diversity, and that study did not publicly release its weights or code. We frame this study’s contribution around the combination of scale, diversity, and open release, as the most extensive such validation of any openly released CMR segmentation model to date.

### Performance at the Basal RVOT/LVOT Level

The basal RVOT/LVOT level is anatomically demanding because the valve planes move through the imaging plane across the cardiac cycle^1^; some approaches track the valve plane on long-axis views to address this^6^, which was not implemented here. Nonetheless, the outflow-tract level was not the weakest-performing position (mean DSC 0.904; §3.3); the apex was substantially more challenging (0.842). Because human inter-observer agreement did not show a comparable apical decline (§3.6), this more likely reflects a genuine model limitation than an unaddressed valve-plane problem at the base.

Figure 3 illustrates representative failure modes, including non-detection of the LV cavity in a case of severe hypertrophic cardiomyopathy, consistent with the known tendency for severe left ventricular hypertrophy to produce multiple, disconnected LV blood-pool regions at peak systole^2^.

### Benchmarking Against Human Inter-observer Variability

Benchmarking automated performance against human inter-observer variability provides a frame of reference largely absent from prior evaluations of automated CMR segmentation tools^3^. Automated LV cavity and right ventricular segmentation matched or exceeded this human benchmark, indicating disagreement of a similar order to that expected between independent human annotators; LV myocardium and ejection-fraction estimation remained below it, identifying these as priority targets for further improvement.

### Interpretation of Clinical Functional Parameter Agreement

The pattern of agreement—strong for volumes/mass, weaker for LVEF, intermediate for RVEF—is consistent with ejection fraction being a ratio of two independently measured volumes, so errors at each phase propagate and can partially compound. LVEF was systematically underestimated (bias −5.40%), whereas RVEF showed the smallest bias despite similarly moderate correlation, suggesting RV volume biases at ED/ES were of comparable magnitude and direction and largely canceled. Absolute errors (MAE 6.20% and 3.97%) fall within ranges reported as clinically acceptable^19^, though inter-observer ICC (0.969, 0.931) still exceeds automated agreement for both.

### Limitations

This study has several limitations. The available repeated-scan data (§3.7) reflect long-term follow-up rather than short-term reproducibility, which was not separately assessed. Training and internal-test data came exclusively from 3.0-T Siemens scanners; multi-vendor/multi-field-strength generalizability, though demonstrated externally, was not itself part of training. Explicit valve-plane tracking was not implemented, and the ischemic heart disease subgroup (n = 5) reflects its source dataset’s composition rather than deliberate sampling. Given the large sample sizes here, statistical significance should be interpreted alongside effect sizes (§3.3), and prospective clinical-impact evaluation remains for future work.

## Conclusion

CorSeg-CineSAX provides a fully automated, openly released framework for short-axis cine CMR segmentation, validated across a substantially larger and more clinically diverse cohort than has previously accompanied an openly released model in this domain, with performance at the anatomically demanding basal outflow-tract level comparable to other anatomical positions. By releasing source code, trained weights, and a standalone graphical application together, this work aims to provide a tool that is both reproducible by researchers and directly usable by clinicians without programming expertise.

## Supporting information

all supplement data

## Data Availability

The raw data that support the findings of this study are available from the corresponding author upon reasonable request.
The model and code are available at https://github.com/RunhaoXu2003/CorSeg

https://www.creatis.insa-lyon.fr/Challenge/acdc/

https://www.ub.edu/mnms/

https://www.kaggle.com/datasets/tailength/m-and-m2-dataset

## Notes

### Competing Interest Statement

The authors have declared no competing interest.

### Author Declarations

Ethics IRB of West China Hospital, Sichuan University gave ethical approval for this work

### Summary of Updates

mainly new feature: segmentation including basal right/left ventricular outflow tract (RVOT/LVOT)

### Funding

- Chengdu Science and Technology Program: 2026-XT00-00016-GX
- Noncommunicable Chronic Diseases-National Science and Technology Major Project: 2024ZD0521802
- Sichuan Health Research: ZH2026-103

