## Supplementary material for "CorSeg-CineSAX: An Open-Source Deep Learning Framework for Fully Automatic Segmentation of Short-Axis Cine Cardiac MRI Across Multiple Cardiac Diseases": all supplement data

### **Supplementary Methods**

### **S1. Data Preprocessing**

All images were stored in NIfTI format. The preprocessing pipeline was applied identically to training and test data and consisted of the following sequential steps.

**Loading and channel formatting:** NIfTI files were loaded and converted to channel-first format (C × H × W); pseudo-3D volumes (single-slice volumes stored as C × H × W × 1) were squeezed to 2D.

**Label remapping:** only the three target foreground labels (LV myocardium = 1, LV cavity = 2, RV = 3) were retained; all other label values were remapped to background (0).

**Affine matrix correction:** to prevent numerical failures during spatial resampling caused by degenerate affine matrices from oblique acquisition orientations, a clean diagonal affine matrix was constructed from the original voxel spacings while preserving the origin translation.

**Spatial resampling:** all images and corresponding segmentation masks were resampled to a uniform in-plane pixel spacing of 1.25 × 1.25 mm, using bilinear interpolation for images and nearest-neighbor interpolation for masks.

**Spatial resizing:** images were resized to a fixed spatial dimension of 224 × 224 pixels via center cropping or zero-padding as necessary.

**Intensity normalization:** per-slice z-score normalization was applied to image intensities, computed exclusively over non-zero voxels; zero-valued background voxels were left unchanged.

The full field-of-view was used as input without any region-of-interest (ROI) cropping, eliminating the need for a separate cardiac localization step and allowing the model to be applied to any CMR short-axis image regardless of field-of-view size or cardiac positioning.

Data augmentation was applied exclusively during training and included both geometric and intensity transformations: random horizontal and vertical flips (probability 0.5 each); random rotation over the full 360° range (probability 0.5); random scaling between 0.85× and 1.15× (probability 0.3); random affine translation up to ±22 pixels and shear up to ±0.05 (probability 0.3); elastic deformation with a grid spacing of 20 pixels and magnitude of 1–2 pixels (probability 0.2); random intensity shift of ±15% (probability 0.3); random intensity scaling of ±15% (probability 0.3); additive Gaussian noise with standard deviation 0.08 (probability 0.2); and random gamma correction with γ ∈ [0.7, 1.5] (probability 0.2). For geometric augmentations, bilinear interpolation was used for images and nearest-neighbor interpolation for masks to preserve label integrity.

### **S2. Model Architecture**

This study employed the MedNeXt^1^ architecture for cardiac structure segmentation — a convolutional network for medical image segmentation inspired by ConvNeXt, with an encoder–decoder structure, skip connections, large convolutional kernels, depthwise separable convolutions, GELU activations, and Layer Normalization in place of Batch Normalization. The Large (L) variant was selected, with a convolutional kernel size of 5 × 5, configured for 2D segmentation with a single input channel (grayscale MRI) and four output channels (background plus three foreground structures). The model was implemented using the Medical Open Network for Artificial Intelligence (MONAI) framework (version 1.5.1)^2^.

A 2D, slice-by-slice segmentation strategy was adopted deliberately, processing each short-axis image independently without 3D volumetric context, temporal sequences, or prior ROI localization. While this sacrifices inter-slice spatial continuity, it maximizes compatibility across diverse clinical scenarios, since the model can process images regardless of data completeness, acquisition protocol, or missing slices/phases.

### **S3. Training Protocol**

The model was trained using the MONAI framework with PyTorch 2.8.0 on an NVIDIA A100 GPU (40 GB memory).

**Loss function:** a composite loss combining Dice loss (computed over the three foreground classes, softmax activation), Focal loss^3^, and weighted cross-entropy loss, with class weights of 0.1/1.0/1.0/1.0 for background/LV myocardium/LV cavity/RV to mitigate class imbalance.

**Optimizer:** AdamW^4^, with a warmup-then-cosine-annealing learning-rate schedule (linear warmup over the first 5 epochs, followed by cosine annealing).

**Training configuration:** maximum 1,000 epochs, batch size 32, mixed-precision (FP16) training via automatic mixed precision with gradient scaling, gradient clipping (maximum norm 1.0), data shuffled each epoch.

**Evaluation during training:** the model was evaluated on the internal test set every 5 epochs and whenever training mean Dice improved by more than 0.005 since the last evaluation; the checkpoint with the highest internal-test mean Dice was retained as the final model.

**Reproducibility:** a global random seed of 42 was set for Python, NumPy, and PyTorch random number generators.

### **Supplementary Figures**

**Figure S1. Qualitative comparison of inter-observer manual segmentation variability.**

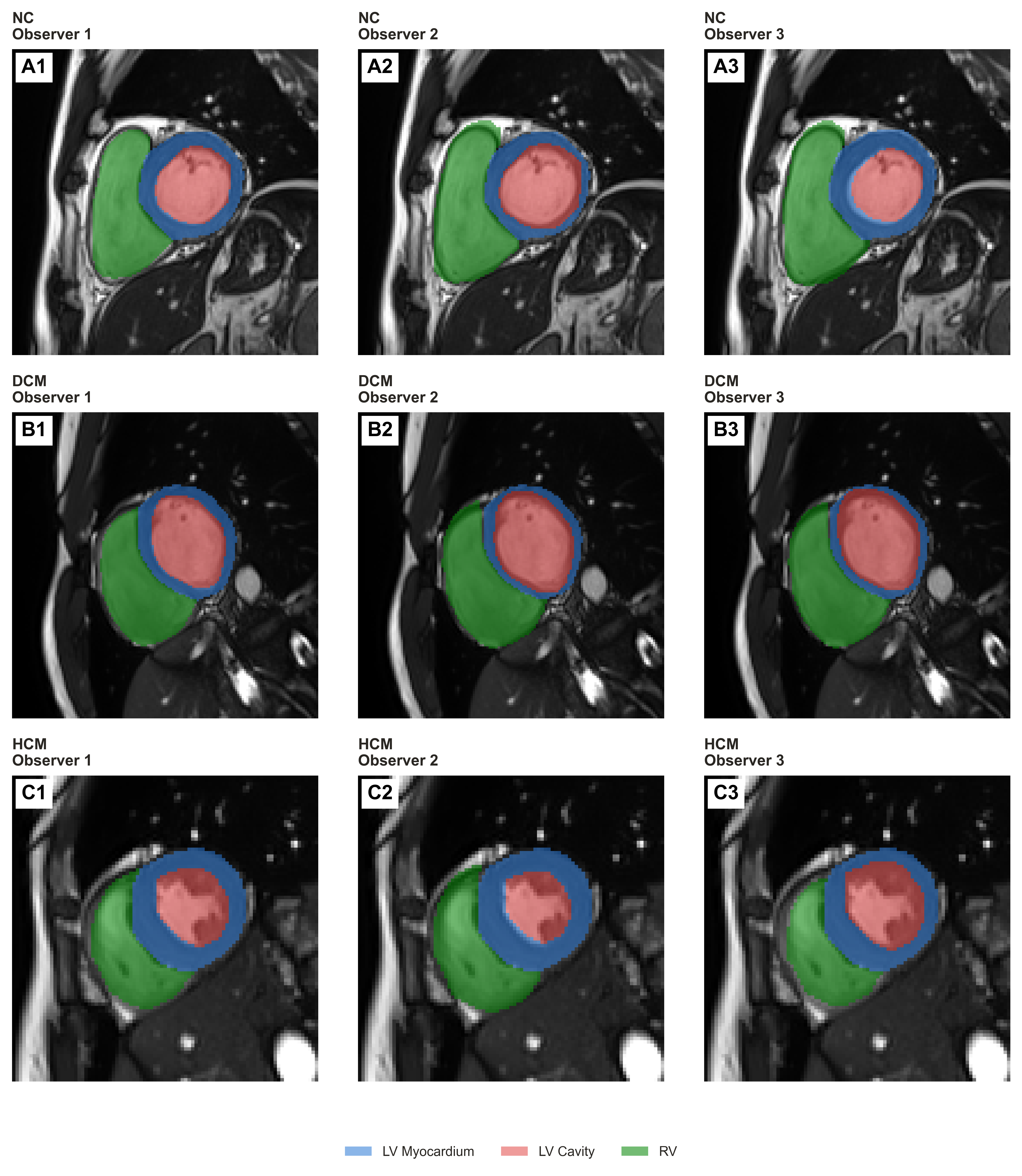

Basal-level masks (blue, LV myocardium; red, LV cavity; green, RV) from three independent observers, one case each of **(A)** NC, **(B)** DCM, **(C)** HCM. Observer 1 = reference standard used as ground truth throughout; Observers 2–3 = independent re-annotators. Same slice per row, illustrating the inter-observer boundary variability quantified in Table S7.

**Figure S2. Atlas of segmentation examples across all 16 disease categories.**

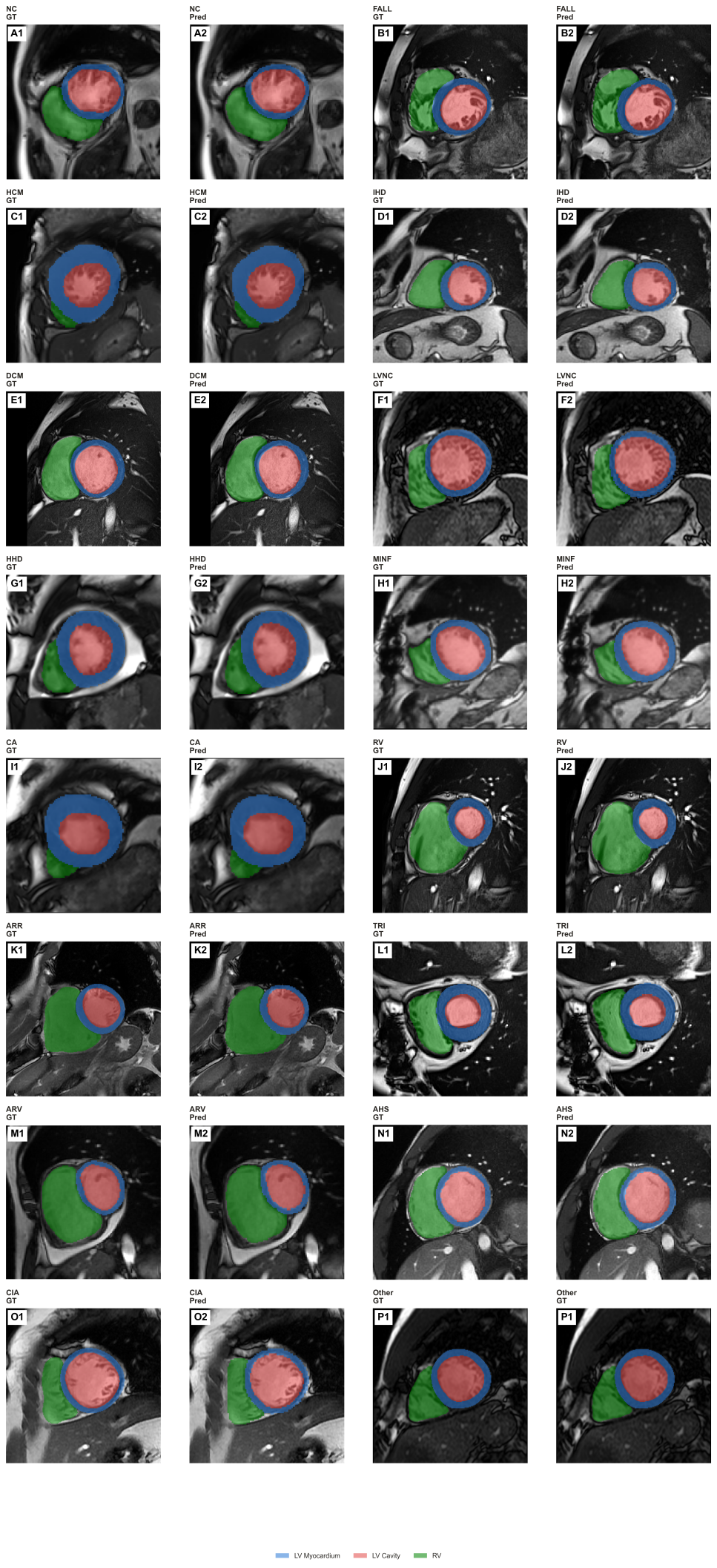

GT (left) and Pred (right) masks for one case each of the 16 disease categories: **(A)** NC; **(B)** Tetralogy of Fallot (FALL); **(C)** HCM; **(D)** ischemic heart disease (IHD); **(E)** DCM; **(F)** LV non-compaction (LVNC); **(G)** HHD; **(H)** myocardial infarction with reduced EF (MINF); **(I)** CA; **(J)** abnormal RV (ACDC/M&Ms2 “RV”); **(K)** arrhythmogenic cardiomyopathy (ARR); **(L)** tricuspid regurgitation (TRI); **(M)** abnormal RV (M&Ms1 “ARV”); **(N)** athlete heart syndrome (AHS); **(O)** inter-atrial communication (CIA); **(P)** unclassified/residual cases (Other). Panels A, C, E, G, I were seen in training; all others were external-only, illustrating cross-disease generalization. J and M represent the same clinical concept under different dataset-specific labels.

**Figure S3. Qualitative segmentation at the basal LVOT, RVOT, and right-ventricular levels: normal control and two pathological examples at each location.**

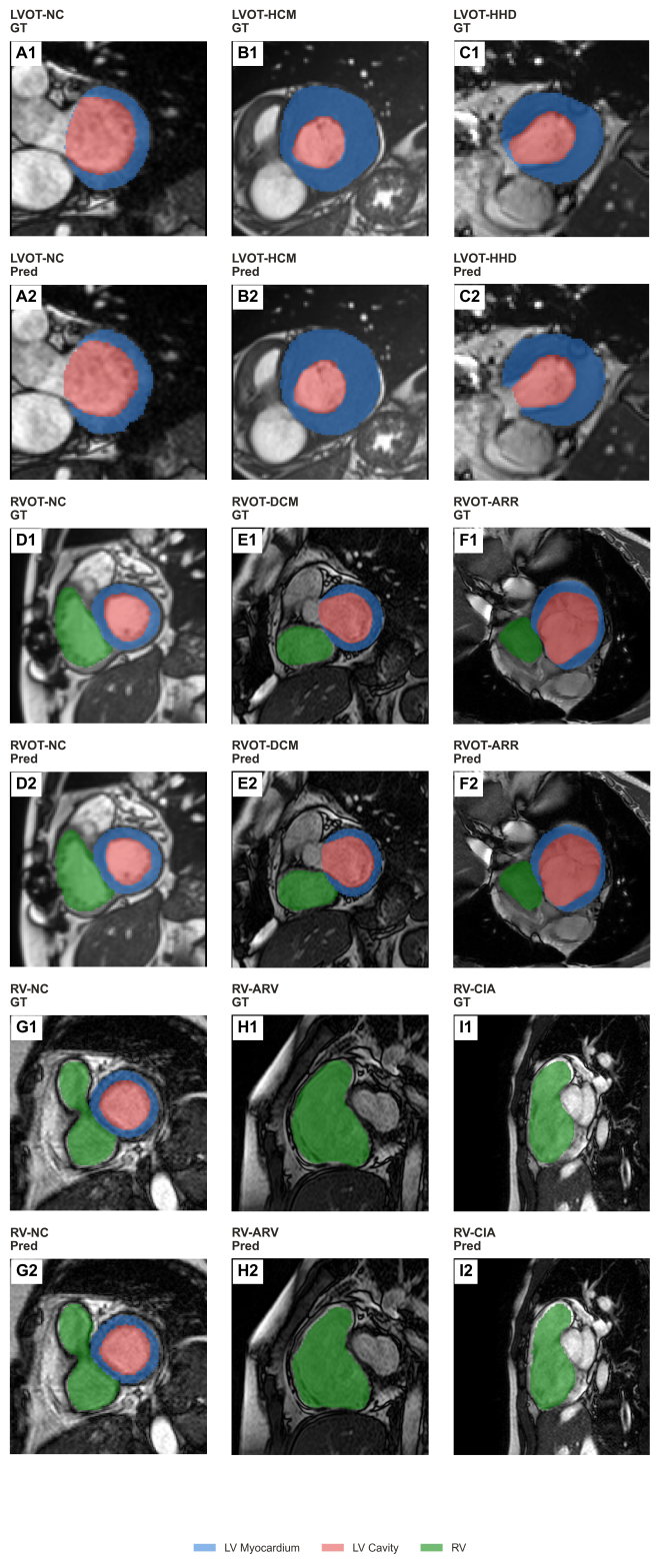

GT (odd-numbered panels) and Pred (even-numbered panels) short-axis masks (blue, LV myocardium; red, LV cavity; green, RV; solid fill; no post-processing) for a normal control and two pathological cases at each of three anatomical locations. (A–C) LVOT level: (A) normal control (NC); (B) hypertrophic cardiomyopathy (HCM); (C) hypertensive heart disease (HHD). (D–F) RVOT level: (D) NC; (E) dilated cardiomyopathy (DCM); (F) arrhythmogenic cardiomyopathy (ARR). (G–I) RV level: (G) NC; (H) abnormal right ventricle (ARV); (I) inter-atrial communication (CIA); in H and I, only the RV was present within the selected slice, so LV myocardium/cavity are not visible. Pathological examples span both disease categories represented in training (HCM, HHD, DCM) and categories present exclusively in the external test datasets (ARR, ARV, CIA).

### **Supplementary Tables**

**Table S1. Segmentation performance stratified by disease category.**

| **label** | **Subjects** | **Slices** | **LVM DSC** | **LVC DSC** | **RV DSC** | **mean DSC** | **HD95 LVM** | **HD95 LVC** | **HD95 RV** |
| --- | --- | --- | --- | --- | --- | --- | --- | --- | --- |
| AHS | 3 | 76 | 0.831±0.053 (0.67–0.91) | 0.915±0.056 (0.74–0.97) | 0.907±0.045 (0.80–0.97) | 0.884 | 3.11±0.49 | 2.91±0.66 | 3.33±0.80 |
| ARR | 35 | 794 | 0.830±0.075 (0.00–0.94) | 0.934±0.038 (0.61–0.98) | 0.914±0.074 (0.00–0.98) | 0.893 | 2.94±1.66 | 2.68±0.85 | 3.15±1.99 |
| ARV | 15 | 356 | 0.820±0.074 (0.51–0.92) | 0.920±0.095 (0.00–0.97) | 0.906±0.086 (0.00–0.98) | 0.882 | 3.14±1.25 | 2.84±0.85 | 4.02±3.35 |
| CA | 72 | 1114 | 0.926±0.033 (0.75–0.98) | 0.946±0.036 (0.60–0.99) | 0.938±0.075 (0.00–0.99) | 0.936 | 2.10±0.76 | 2.34±1.25 | 2.29±2.78 |
| CIA | 35 | 788 | 0.822±0.068 (0.37–0.95) | 0.929±0.062 (0.00–0.98) | 0.927±0.053 (0.44–0.98) | 0.893 | 2.59±1.05 | 2.38±0.85 | 3.33±2.48 |
| DCM | 187 | 4376 | 0.822±0.072 (0.00–0.96) | 0.933±0.069 (0.00–0.98) | 0.895±0.096 (0.00–0.98) | 0.883 | 2.94±1.65 | 2.68±0.94 | 3.33±3.56 |
| FALL | 35 | 774 | 0.840±0.088 (0.00–0.94) | 0.941±0.037 (0.58–0.98) | 0.930±0.087 (0.00–0.98) | 0.904 | 2.74±2.55 | 2.47±2.79 | 3.20±2.32 |
| HCM | 289 | 5592 | 0.890±0.066 (0.00–0.99) | 0.919±0.079 (0.00–0.99) | 0.904±0.116 (0.00–0.99) | 0.904 | 2.79±3.36 | 2.60±3.18 | 2.81±2.91 |
| HHD | 116 | 2146 | 0.905±0.053 (0.54–0.99) | 0.941±0.053 (0.00–0.99) | 0.939±0.068 (0.00–1.00) | 0.928 | 2.38±3.56 | 2.51±3.70 | 2.16±2.40 |
| IHD | 5 | 126 | 0.834±0.067 (0.57–0.93) | 0.916±0.057 (0.57–0.96) | 0.889±0.072 (0.58–0.97) | 0.880 | 2.89±0.52 | 2.87±0.68 | 3.23±1.01 |
| LVNC | 2 | 50 | 0.812±0.059 (0.69–0.89) | 0.940±0.036 (0.80–0.97) | 0.915±0.025 (0.85–0.96) | 0.889 | 2.57±0.49 | 2.32±0.40 | 2.85±1.16 |
| MINF | 30 | 534 | 0.829±0.079 (0.00–0.94) | 0.927±0.090 (0.00–0.98) | 0.867±0.125 (0.00–0.97) | 0.874 | 2.15±0.66 | 2.01±0.58 | 2.66±3.83 |
| NC | 566 | 11878 | 0.823±0.086 (0.00–0.95) | 0.911±0.078 (0.00–0.98) | 0.910±0.079 (0.00–0.98) | 0.881 | 2.88±3.79 | 2.83±4.05 | 2.98±2.23 |
| Other | 25 | 650 | 0.829±0.080 (0.00–0.95) | 0.916±0.094 (0.00–0.98) | 0.891±0.124 (0.00–0.97) | 0.879 | 2.82±0.78 | 2.64±0.75 | 3.36±3.05 |
| RV | 60 | 1522 | 0.823±0.072 (0.17–0.94) | 0.921±0.079 (0.00–0.98) | 0.913±0.096 (0.00–0.98) | 0.886 | 2.33±1.25 | 2.15±1.09 | 2.72±2.18 |
| TRI | 30 | 664 | 0.808±0.079 (0.35–0.94) | 0.927±0.060 (0.00–0.98) | 0.911±0.081 (0.00–0.97) | 0.882 | 3.13±1.69 | 2.90±1.26 | 3.24±2.16 |

*P value (not shown in table above): Comparison across all disease categories (subject-level mean DSC): Kruskal-Wallis test, H = 432.7, P = 1.6×10⁻⁸³ (Holm-Bonferroni–adjusted P = 4.9×10⁻⁸³), ε² = 0.28 (n = 1503 subjects).*

DSC and HD95 (mm), mean ± SD (min–max), by disease category (combined test set). CA/DCM/HCM/HHD/NC were seen in training; the remaining 11 categories (AHS, ARR, ARV, CIA, FALL, IHD, LVNC, MINF, RV, TRI, Other) were external-only. Abbreviations: AHS, athlete heart syndrome; ARR, arrhythmogenic cardiomyopathy; ARV, abnormal right ventricle (M&Ms1); CIA, inter-atrial communication; FALL, Tetralogy of Fallot; IHD, ischemic heart disease; LVNC, LV non-compaction; MINF, myocardial infarction with reduced EF; RV, abnormal/dilated right ventricle (ACDC/M&Ms2, distinct from ARV owing to source-dataset labelling); TRI, tricuspid regurgitation; Other, unclassified residual cases. IHD (n = 5) reflects its sole source dataset (ACDC).

**Table S2. Segmentation performance stratified by imaging center.**

| **center** | **Subjects** | **Slices** | **LVM DSC** | **LVC DSC** | **RV DSC** | **mean DSC** | **HD95 LVM** | **HD95 LVC** | **HD95 RV** |
| --- | --- | --- | --- | --- | --- | --- | --- | --- | --- |
| Anqing Municipal Hospital | 104 | 1896 | 0.833±0.079 (0.00–0.95) | 0.913±0.074 (0.00–0.98) | 0.919±0.060 (0.00–0.98) | 0.889 | 2.96±6.88 | 3.11±8.41 | 2.78±1.84 |
| Beijing Hospital | 67 | 1282 | 0.790±0.100 (0.00–0.93) | 0.882±0.100 (0.00–0.97) | 0.905±0.076 (0.00–0.97) | 0.859 | 3.54±1.90 | 3.58±1.37 | 3.55±1.03 |
| Clinica Creu Blanca | 50 | 1266 | 0.824±0.086 (0.00–0.95) | 0.924±0.067 (0.00–0.98) | 0.890±0.097 (0.00–0.97) | 0.879 | 2.79±2.86 | 2.54±0.82 | 3.47±3.15 |
| Clinica Sagrada Familia | 162 | 3274 | 0.830±0.075 (0.00–0.95) | 0.912±0.073 (0.00–0.98) | 0.896±0.099 (0.00–0.97) | 0.880 | 3.03±1.20 | 2.84±1.07 | 3.45±3.21 |
| Fujian Medical University Union Hospital | 106 | 2748 | 0.823±0.113 (0.00–0.95) | 0.906±0.096 (0.00–0.98) | 0.910±0.083 (0.00–0.98) | 0.879 | 3.05±4.95 | 2.96±4.24 | 2.86±1.39 |
| Handan Central Hospital | 95 | 1820 | 0.841±0.061 (0.53–0.95) | 0.924±0.049 (0.55–0.98) | 0.925±0.063 (0.00–0.98) | 0.896 | 2.60±0.65 | 2.63±1.30 | 2.78±1.01 |
| Hospital Universitari Dexeus | 128 | 3064 | 0.827±0.074 (0.34–0.95) | 0.917±0.075 (0.00–0.98) | 0.896±0.090 (0.00–0.98) | 0.880 | 2.91±1.10 | 2.75±1.04 | 3.27±2.52 |
| Hospital Vall'd Hebron | 314 | 7268 | 0.832±0.072 (0.00–0.96) | 0.932±0.065 (0.00–0.98) | 0.909±0.096 (0.00–0.98) | 0.891 | 3.06±1.88 | 2.69±1.31 | 3.35±2.88 |
| The First Affiliated Hospital of Anhui Medical University | 278 | 4680 | 0.928±0.035 (0.21–0.99) | 0.944±0.040 (0.25–0.99) | 0.940±0.078 (0.00–1.00) | 0.937 | 2.22±3.96 | 2.37±3.97 | 2.10±2.77 |
| Universitatsklinikum | 51 | 1164 | 0.817±0.082 (0.00–0.96) | 0.910±0.091 (0.00–0.98) | 0.894±0.083 (0.00–0.97) | 0.874 | 3.07±0.85 | 2.94±0.77 | 3.50±2.42 |
| University Hospital of Dijon | 150 | 2978 | 0.838±0.069 (0.00–0.96) | 0.914±0.094 (0.00–0.98) | 0.887±0.118 (0.00–0.98) | 0.880 | 2.08±0.73 | 1.93±0.62 | 2.45±3.26 |

*P value (not shown in table above): Comparison across all imaging centers (subject-level mean DSC): Kruskal-Wallis test, H = 798.1, P = 5.4×10⁻¹⁶⁵ (Holm-Bonferroni–adjusted P = 2.7×10⁻¹⁶⁴), ε² = 0.53 (n = 1505 subjects).*

DSC and HD95 (mm), mean ± SD (min–max), by imaging center (11 test-set centers; training-only centers not shown).

**Table S3a. Segmentation performance stratified by scanner vendor.**

| **vendor_norm** | **Subjects** | **Slices** | **LVM DSC** | **LVC DSC** | **RV DSC** | **mean DSC** | **HD95 LVM** | **HD95 LVC** | **HD95 RV** |
| --- | --- | --- | --- | --- | --- | --- | --- | --- | --- |
| Canon | 75 | 1788 | 0.826±0.079 (0.34–0.95) | 0.909±0.079 (0.00–0.98) | 0.888±0.105 (0.00–0.98) | 0.874 | 2.99±0.89 | 2.85±0.79 | 3.29±2.71 |
| GE | 104 | 2440 | 0.823±0.075 (0.00–0.96) | 0.919±0.081 (0.00–0.98) | 0.901±0.075 (0.00–0.98) | 0.881 | 2.93±1.12 | 2.78±1.07 | 3.37±2.35 |
| Philips | 212 | 4540 | 0.828±0.078 (0.00–0.95) | 0.915±0.072 (0.00–0.98) | 0.895±0.098 (0.00–0.97) | 0.879 | 2.97±1.76 | 2.77±1.02 | 3.46±3.19 |
| Siemens | 1114 | 22672 | 0.855±0.083 (0.00–0.99) | 0.925±0.072 (0.00–0.99) | 0.916±0.090 (0.00–1.00) | 0.899 | 2.68±3.39 | 2.59±3.50 | 2.75±2.60 |

*P value (not shown in table above): Comparison across scanner vendors (subject-level mean DSC): Kruskal-Wallis test, H = 139.8, P = 4.1×10⁻³⁰ (Holm-Bonferroni–adjusted P = 4.1×10⁻³⁰), ε² = 0.09 (n = 1505 subjects).*

DSC and HD95 (mm), mean ± SD (min–max), by scanner vendor (Siemens, Philips, GE, Canon), combined test set.

**Table S3b. Segmentation performance stratified by magnetic field strength.**

| **field_strength** | **Subjects** | **Slices** | **LVM DSC** | **LVC DSC** | **RV DSC** | **mean DSC** | **HD95 LVM** | **HD95 LVC** | **HD95 RV** |
| --- | --- | --- | --- | --- | --- | --- | --- | --- | --- |
| 1.5 | 593 | 13396 | 0.827±0.077 (0.00–0.96) | 0.922±0.070 (0.00–0.98) | 0.902±0.095 (0.00–0.98) | 0.884 | 3.00±1.72 | 2.74±1.16 | 3.30±2.68 |
| 1.5/3.0 | 150 | 2978 | 0.838±0.069 (0.00–0.96) | 0.914±0.094 (0.00–0.98) | 0.887±0.118 (0.00–0.98) | 0.880 | 2.08±0.73 | 1.93±0.62 | 2.45±3.26 |
| 3.0 | 762 | 15066 | 0.866±0.086 (0.00–0.99) | 0.925±0.071 (0.00–0.99) | 0.922±0.080 (0.00–1.00) | 0.905 | 2.70±4.01 | 2.73±4.22 | 2.73±2.51 |

*P value (not shown in table above): Comparison across field strengths (subject-level mean DSC): Kruskal-Wallis test, H = 189.0, P = 9.2×10⁻⁴² (Holm-Bonferroni–adjusted P = 1.8×10⁻⁴¹), ε² = 0.12 (n = 1505 subjects).*

DSC and HD95 (mm), mean ± SD (min–max), by field strength (1.5 T, 3.0 T, and “1.5/3.0 T” for ACDC, which lacks per-subject field-strength data), combined test set.

**Table S4. Segmentation performance stratified by cardiac phase.**

| **phase_type** | **Subjects** | **Slices** | **LVM DSC** | **LVC DSC** | **RV DSC** | **mean DSC** | **HD95 LVM** | **HD95 LVC** | **HD95 RV** |
| --- | --- | --- | --- | --- | --- | --- | --- | --- | --- |
| ED | 1505 | 15720 | 0.822±0.080 (0.00–0.98) | 0.940±0.059 (0.00–0.99) | 0.924±0.079 (0.00–0.99) | 0.895 | 2.59±2.23 | 2.54±2.05 | 2.80±2.38 |
| ES | 1505 | 15720 | 0.876±0.075 (0.00–0.99) | 0.903±0.083 (0.00–0.99) | 0.893±0.103 (0.00–1.00) | 0.891 | 2.94±3.71 | 2.76±3.92 | 3.06±3.04 |

*P value (not shown in table above): Comparison between ED and ES (subject-level mean DSC, paired): Wilcoxon signed-rank test, W = 457641, P = 1.0×10⁻¹⁰ (medians 0.891 vs. 0.888; n = 1505 subjects).*

DSC and HD95 (mm), mean ± SD (min–max), for ED vs. ES, combined test set.

**Table S5. Segmentation performance stratified by source dataset.**

| **dataset** | **Subjects** | **Slices** | **LVM DSC** | **LVC DSC** | **RV DSC** | **mean DSC** | **HD95 LVM** | **HD95 LVC** | **HD95 RV** |
| --- | --- | --- | --- | --- | --- | --- | --- | --- | --- |
| Data-ACDC | 150 | 2978 | 0.838±0.069 (0.00–0.96) | 0.914±0.094 (0.00–0.98) | 0.887±0.118 (0.00–0.98) | 0.880 | 2.08±0.73 | 1.93±0.62 | 2.45±3.26 |
| Data-AnhuiCA | 72 | 1114 | 0.926±0.033 (0.75–0.98) | 0.946±0.036 (0.60–0.99) | 0.938±0.075 (0.00–0.99) | 0.936 | 2.10±0.76 | 2.34±1.25 | 2.29±2.78 |
| Data-AnhuiHCM | 114 | 1956 | 0.935±0.035 (0.21–0.99) | 0.940±0.046 (0.25–0.99) | 0.934±0.092 (0.00–0.99) | 0.936 | 2.29±4.95 | 2.35±4.82 | 2.12±2.98 |
| Data-AnhuiHHD | 92 | 1610 | 0.920±0.035 (0.54–0.99) | 0.949±0.036 (0.41–0.99) | 0.948±0.060 (0.00–1.00) | 0.939 | 2.22±3.92 | 2.42±4.09 | 1.95±2.49 |
| Data-MnM1 | 345 | 7908 | 0.828±0.076 (0.00–0.96) | 0.914±0.083 (0.00–0.98) | 0.889±0.103 (0.00–0.98) | 0.877 | 3.06±1.50 | 2.83±0.89 | 3.60±3.25 |
| Data-MnM2 | 360 | 8128 | 0.829±0.074 (0.00–0.96) | 0.931±0.056 (0.00–0.98) | 0.913±0.085 (0.00–0.98) | 0.891 | 2.96±1.81 | 2.66±1.35 | 3.16±2.46 |
| Data-MultiCenterNC | 372 | 7746 | 0.825±0.092 (0.00–0.95) | 0.909±0.082 (0.00–0.98) | 0.916±0.072 (0.00–0.98) | 0.883 | 2.98±4.59 | 3.01±5.02 | 2.91±1.42 |

*P value (not shown in table above): Comparison across source datasets (subject-level mean DSC): Kruskal-Wallis test, H = 700.6, P = 4.5×10⁻¹⁴⁸ (Holm-Bonferroni–adjusted P = 1.8×10⁻¹⁴⁷), ε² = 0.46 (n = 1505 subjects).*

DSC and HD95 (mm), mean ± SD (min–max), by source dataset: ACDC; private AnhuiCA/AnhuiHCM/AnhuiHHD; M&Ms1; M&Ms2; private multi-center NC cohort.

**Table S6. Long-term repeat-scan reproducibility (test-retest).**

| **Param** | **n_scans** | **ICC(A,1)** | **Within-subject CoV%** | **mean±sd** |
| --- | --- | --- | --- | --- |
| LVEDV | 47 | 0.369 | 25.9 | 146.3±87.0 |
| LVESV | 47 | 0.338 | 34.7 | 77.3±64.1 |
| LVEF | 47 | 0.412 | 14.5 | 50.4±13.4 |
| LVMass | 47 | 0.456 | 22.6 | 130.1±70.5 |
| RVEDV | 47 | 0.432 | 27.8 | 123.3±64.8 |
| RVESV | 47 | 0.436 | 31.7 | 91.1±53.8 |
| RVEF | 47 | 0.438 | 29.6 | 27.4±13.3 |

ICC(A,1) and within-subject CoV% for 18 subjects, 47 repeat scans (median interval, 376 days; range, 28–1190 days), training/validation cohort only. Reflects long-term physiological change plus measurement variability, not short-term reproducibility.

**Table S7a. Inter-observer variability of manual segmentation by Disease.**

| **Group** | **Subjects** | **Slice-pair comparisons** | **LVM DSC** | **LVC DSC** | **RV DSC** | **mean DSC** |
| --- | --- | --- | --- | --- | --- | --- |
| Overall | 50 | 1306 | 0.875±0.084 (0.27–0.99) | 0.894±0.049 (0.76–1.00) | 0.906±0.048 (0.15–0.99) | 0.892 |
| CA | 10 | 168 | 0.897±0.052 (0.64–0.98) | 0.893±0.048 (0.77–1.00) | 0.900±0.045 (0.68–0.98) | 0.897 |
| DCM | 10 | 250 | 0.819±0.127 (0.27–0.98) | 0.891±0.048 (0.77–0.99) | 0.915±0.037 (0.79–0.99) | 0.875 |
| HCM | 10 | 296 | 0.894±0.062 (0.54–0.99) | 0.892±0.048 (0.77–1.00) | 0.897±0.066 (0.15–0.99) | 0.894 |
| HHD | 10 | 156 | 0.886±0.064 (0.56–0.99) | 0.896±0.049 (0.76–1.00) | 0.907±0.044 (0.75–0.99) | 0.896 |
| NC | 10 | 436 | 0.873±0.075 (0.54–0.99) | 0.897±0.049 (0.77–1.00) | 0.911±0.038 (0.77–0.98) | 0.894 |

Pairwise DSC (mean ± SD, min–max) among three independent observers (Observer 1 = reference standard; Observers 2–3 = independent re-annotators), 50 subjects (10 per disease × ED/ES), by disease category.

**Table S7b. Inter-observer variability of manual segmentation by Slice Position.**

| **Group** | **Subjects** | **Slice-pair comparisons** | **LVM DSC** | **LVC DSC** | **RV DSC** | **mean DSC** |
| --- | --- | --- | --- | --- | --- | --- |
| basal_outflow | 49 | 236 | 0.884±0.080 (0.36–0.99) | 0.895±0.049 (0.77–1.00) | 0.912±0.040 (0.75–0.98) | 0.897 |
| basal | 49 | 398 | 0.866±0.088 (0.27–0.99) | 0.895±0.048 (0.76–0.99) | 0.909±0.039 (0.75–0.99) | 0.890 |
| mid | 49 | 352 | 0.870±0.091 (0.27–0.99) | 0.893±0.049 (0.77–0.99) | 0.904±0.048 (0.59–0.99) | 0.889 |
| apical | 49 | 304 | 0.892±0.061 (0.54–0.99) | 0.892±0.048 (0.77–1.00) | 0.895±0.082 (0.15–0.98) | 0.893 |

Pairwise DSC among the same three observers, by anatomical slice position; comparable with the basal-outflow-tract row in Table 3.

**Table S7c. Inter-observer variability of functional parameters.**

| **Param** | **n** | **ICC(A,1) (3 raters)** | **Observer_1** | **Observer_2** | **Observer_3** |
| --- | --- | --- | --- | --- | --- |
| LVEDV | 49 | 0.985 | 144.5±63.4 | 148.9±68.5 | 149.2±68.3 |
| LVESV | 49 | 0.987 | 61.4±42.6 | 63.4±45.0 | 63.0±44.9 |
| LVEF | 49 | 0.969 | 58.4±19.2 | 58.1±20.3 | 58.5±19.3 |
| LVMass | 49 | 0.983 | 132.6±65.6 | 128.7±63.6 | 128.1±61.8 |
| RVEDV | 49 | 0.994 | 128.5±71.6 | 127.8±72.2 | 128.7±71.5 |
| RVESV | 49 | 0.989 | 71.8±36.7 | 70.8±37.4 | 69.2±35.3 |
| RVEF | 49 | 0.931 | 41.7±15.6 | 42.0±15.8 | 43.7±16.1 |

Three-rater ICC(A,1) for each functional parameter among the same three observers (n = 49–50); comparable with Table 4.
